# The complex relationship of air pollution and neighborhood socioeconomic deprivation and their association with cognitive decline

**DOI:** 10.1101/2022.03.29.22273134

**Authors:** Grace Christensen, Zhenjiang Li, John Pearce, Michele Marcus, James J. Lah, Lance A. Waller, Stefanie Ebelt, Anke Huels

## Abstract

**Background:** Air pollution and neighborhood socioeconomic status (nSES) have been shown to affect cognitive decline in older adults. In previous studies, nSES acts as both a confounder and an effect modifier between air pollution and cognitive decline.

**Objectives:** This study aims to examine the individual and joint effects of air pollution and nSES on cognitive decline on adults 50 years and older in Metro Atlanta, USA.

**Methods:** Perceived memory and cognitive decline was assessed in 11,897 participants aged 50+ years from the Emory Healthy Aging Study (EHAS) using the cognitive function instrument (CFI). Three-year average air pollution concentrations for 12 pollutants and 16 nSES characteristics were matched to participants using census tracts. Individual exposure linear regression and LASSO models explore individual exposure effects. Environmental mixture modeling methods including, self-organizing maps (SOM), Bayesian kernel machine regression (BKMR), and quantile-based G-computation explore joint effects, and effect modification between air pollutants and nSES characteristics on cognitive decline.

**Results:** Participants living in areas with higher air pollution concentrations and lower nSES experienced higher CFI scores (beta: 0.121; 95% CI: 0.076, 0.167) compared to participants living in areas with low air pollution and high nSES. Additionally, the BKMR model showed a significant overall mixture effect on cognitive decline, indicating synergy between air pollution and nSES. These joint effects explain protective effects observed in single-pollutant linear regression models, even after adjustment for confounding by nSES (e.g., an IQR increase in CO was associated with a 0.038-point decrease (95% CI: -0.06, -0.01) in CFI score).

**Discussion:** Observed protective effects of single air pollutants on cognitive decline can be explained by joint effects and effect modification of air pollutants and nSES. Researchers must consider nSES as an effect modifier if not a co-exposure to better understand the complex relationships between air pollution and nSES in urban settings.

## Introduction

Exposure to ambient air pollution, particularly fine particulate matter (PM_2.5_), in adulthood has been associated with cognitive decline and dementias like Alzheimer’s disease (Paul et al., 2019; Peters et al., 2019; Power et al., 2016; Weuve et al., 2021). Animal models suggest that air pollution affects the central nervous system through pathways involving inflammation and oxidative stress (Costa et al., 2020; Hahad et al., 2020). Though exposure to air pollutants occurs as a mixture of correlated pollutants, it is not treated as such in most epidemiological studies, including those that have assessed cognitive decline, which often rely on the use of single pollutant models (Dominici et al., 2010). Consideration of neighborhood socioeconomic status (nSES) is also important to understand the full impact of air pollution on cognitive decline.

It is well known that individuals living in disadvantaged neighborhoods are often exposed to higher concentrations of air pollution than individuals living in more socioeconomically advantaged neighborhoods (Landrigan et al., 2018; Maantay, 2002). Additionally, air pollution and social vulnerability are hypothesized to interact leading to impaired health outcomes, including cognitive decline (J. A. Ailshire & Clarke, 2015). Social stressors that occur in disadvantaged neighborhoods are an integral part of the ‘triple jeopardy’ of environmental injustice. The triple jeopardy hypothesis examines how low nSES communities experience higher exposure to air pollution and increased susceptibility to poor health due to increased psychosocial stressors among other factors, resulting in health disparities (Hajat et al., 2015). Exposure to both neighborhood social stressors and air pollution may jointly affect cognitive decline in older adults through various biological pathways, but these interactions are poorly understood. Toxicology studies suggest that social stressors may lower the brain’s threshold for neurotoxicity, thus making those living in disadvantaged neighborhoods more vulnerable to the harmful effects of air pollution (Lupien et al., 2009; McEwen & Tucker, 2011).

Despite the importance and relevance of the concept of environmental injustice for cognitive health, few epidemiologic studies on air pollution and cognition have considered nSES as a confounder (J. A. Ailshire & Crimmins, 2014; Bowe et al., 2019; Cullen et al., 2018; Li et al., 2021) or effect modifier (J. Ailshire et al., 2017; Li et al., 2021). One study treated nSES as an effect modifier and found the association between PM_2.5_ and cognitive errors was stronger in older adults living in high stress neighborhoods (J. Ailshire et al., 2017). Another study found that nSES was both a confounder and an effect modifier of the association of air pollution and cognitive decline (Li et al., 2021). These studies show that it is imperative to include socioeconomic context of participants in studies of air pollution and cognitive decline. One methodologic challenge of analyses related to environmental injustice is the high correlation between environmental and social stressors. Previous studies have evaluated interactions and effect modifications of air pollutants and nSES by including interaction terms or conducting stratified linear regression models (J. Ailshire et al., 2017; Li et al., 2021). Modeling environmental exposures, including pollutants and socioeconomic contexts of the population, as a mixture is a necessary next step to describe the associations between environmental pollutants and health effects and understand their joint and potentially synergistic effects (Carlin et al., 2013; Taylor et al., 2016).

Recently, statistical methods to evaluate individual, joint, and interaction effects of environmental mixtures have been developed (Bobb et al., 2015; Carlin et al., 2013; Keil et al., 2020; Pearce et al., 2016). These methods all evaluate the complex problem of environmental mixtures. However, since they each address slightly different aspects of how mixtures affect health, multiple methods must be used for a holistic assessment of the mixture effects, particularly in diverse urban settings. Mixture methods developed to investigate individual effects of highly correlated exposure variables include Bayesian Kernel Machine Regression (BKMR), Least Absolute Shrinkage and Selection Operator (LASSO) and quantile-based G-computation. BKMR and quantile-based G-computation further allow estimation of the joint effects of the mixture as a whole and BKMR can estimate interactions between a limited number of exposure mixtures. Self-organizing maps (SOM) utilize a different strategy for analysis of joint effects by identifying pertinent types of multipollutant combinations that jointly affect health. However, the application of these mixture methods has been limited to environmental exposures and none of these methods have been used to quantify the complex relationship between air pollution and nSES in the context of environmental injustice on cognitive decline.

This study aims to use these mixture methods to 1) identify the most harmful air pollutants and nSES indicators in relation to cognitive decline; 2) estimate the joint effects of air pollution and nSES on cognitive decline; and 3) estimate effect modification of air pollutants and nSES characteristics on cognitive decline in adults 50 years and older in Atlanta, GA.

## Methods

### Study Population

The Emory Healthy Aging Study (EHAS) is a large gerontology-based ongoing research study focusing on diseases of older adults, starting in 2015. Enrollment is open to anyone over age 18, living in the US, and sufficiently fluent in English. Recruitment in the Metro-Atlanta area focused on individuals receiving health services at Emory Healthcare as well as their spouses, family members, and associated non-relatives. Enrollment, consent, and all questionnaires were completed online and described elsewhere (Goetz et al., 2019). EHAS participants that enrolled during the 2015 – 2020 period, were 50 years and older at baseline, and living in the Metro-Atlanta area were included in this analysis. The EHAS includes a Health History Questionnaire at enrollment where participants are asked demographic questions, such as age and self-reported race/ethnicity, and health information as well as an assessment of perceived memory and cognitive decline. All participants complete an online consent process prior to enrollment, and the study was approved by the Emory University Institutional Review Board.

### Exposure Assessment

Average ambient air pollution concentrations for the 2008-2010 period were derived from the Community Multiscale Air Quality (CMAQ) chemical transport model for twelve pollutants, Nitrogen Oxides (NO_x_), nitrogen Dioxide (NO_2_), Nitrate (NO_3_), Sulfur Dioxide (SO_2_), Ozone (O_3_), carbon Monoxide (CO), Ammonium (NH_4_), Particulate matter with a diameter 10 microns or less and 2.5 microns or less (PM_10_, PM_2.5_), Sulfate (SO_4_), Elemental Carbon (EC), and Organic Carbon (OC) at a grid resolution of 4-kilometers (Senthilkumar et al., 2019). For the main analysis, participants were assigned CMAQ pollutant concentrations based on their census tract of residence. Specifically, the center of each CMAQ grid cell was geospatially matched to the closest census tract, which was then connected to each participant based on their residential street address at enrollment. Exposures assigned based on residential census tract, as opposed to street address, were used in the main analysis to facilitate the application of certain mixture methods (i.e., Self-Organizing Maps, which require all inputs (i.e., air pollution and nSES) to be at the same spatial resolution) and thus to facilitate comparison of results across mixture methods. In a sensitivity analysis, CMAQ grid cell exposures were assigned to participants based on their residential street address to ensure that there was no bias caused by the broader census tract matching.

Neighborhood socioeconomic characteristics (nSES) were obtained for each Metro-Atlanta census tract from United States Census Bureau’s American Community Survey for the years 2013-2018. The 5-year average estimates were obtained through the R package *tidycensus*. Sixteen nSES indicators representing the six domains poverty/income, racial composition, education, employment, occupation, housing properties, were chosen to represent a mixture of exposures to nSES (Messer et al., 2006). The nSES characteristic median home value was multiplied by -1 to be in line with the other nSES characteristics, meaning that an increasing value indicates lower nSES. Additionally, an indicator for residential stability (the percentage of households that moved into their current residence before 2010) was used to control for confounding due to residential mobility in analyses.

### Assessment of Cognitive Decline

Cognitive decline was measured using the cognitive function instrument (CFI), where higher scores indicate increased perceived memory decline and cognitive decline (Amariglio et al., 2015). The CFI was self-administered and consists of 14 questions that probe subjective cognitive concerns occurring in daily life of older adults. The CFI score is predictive of cognitive decline in older adults, and can reliably track early changes in cognitive function in patients without clinical impairment. Subjective experiences of cognitive decline occur at the late stage of the preclinical, cognitively unimpaired phase of the Alzheimer’s disease continuum and are therefore often considered as the first symptom of dementia (Jack et al., 2018). Unlike other methods to assess cognitive impairment, the CFI does not require an in-person interview and review by a physician (Amariglio et al., 2015). Total CFI score was calculated by scoring the responses to each question (Yes = 1, No = 0, and Maybe = 0.5) to create an instrument ranging from 0 to 14, where higher scores indicate a higher degree of cognitive decline. Total CFI score was right skewed in our sample, and was therefore log-transformed for all analyses.

### Statistical Analysis

To model individual and joint effects of air pollution exposures and nSES characteristics multiple exposure mixture modeling techniques were used. Statistical approaches were selected based on the research question (e.g. individual effect, effect modification, joint effect), and at this time there is no one method appropriate for all exposure mixture research (Taylor et al., 2016). Furthermore, analyses with multiple methods can allow researchers to examine the relationships between environmental mixtures and health outcomes from different perspectives to come to a more comprehensive understanding of the relationships under study.

A directed acyclic graph (DAG) was used to select confounders for the effect of air pollution and nSES on cognitive decline (Figure S1). Potential confounders under consideration included individual age, race/ethnicity, education, and residential stability of the census tract. These potential confounders were used in all adjusted analyses. For the assessment of effects of individual air pollutants, models were additionally adjusted for nSES. As most of the 16 nSES variables are highly correlated (Figure S2B), principal components (PCs) were derived from nSES characteristics that account for 80% of the total variance in nSES. The uncorrelated nSES variables were then included as confounding variables in the association analyses, following the work in Li et al., (2021). For the assessment of joint effects, air pollution and nSES were considered as co-exposures.

### Individual Effects

First, we considered individual linear regression models for each air pollutant and nSES characteristic adjusted for individual age, race/ethnicity, education, and residential stability of the census tract. Associations with air pollution exposures were additionally adjusted for PCs of nSES characteristics. These single-exposure models illustrated how each exposure associates with cognitive decline when not accounting for other co-exposures.

Second, using the R package *glmnet*, multi-pollutant models via LASSO regression were conducted to determine which mixture components (16 nSES and 12 air pollutants) most contribute to the mixture effect on the natural log of CFI (ln(CFI)), while accounting for the other co-exposures. LASSO regression is a supervised regression analysis method that selects variables most influential on the outcome and shrinks the effect estimates of the variables that are not influential on the outcome (Tibshirani, 1996). This method performs variable selection while controlling for co-exposures and confounding.

BKMR estimates individual effects of each of the exposures and determines which mixture components contribute to the mixture’s effect on the outcome, while accounting for the other exposures in the mixture (Bobb et al., 2018). BKMR uses a specified kernel function to model the exposure mixture’s effect on an outcome, adjusting for confounders. This method allows for non-linear relationships between exposure and outcome, and can additionally model both the total mixture effect and the individual effect of each component accounting for collinearity (Bobb et al., 2015, 2018). BKMR analyses were done using the *bkmr* package in R (ver. 3.6.1). BKMR results are based on 20,000 iterations and were adjusted for confounders as described above.

Quantile-based G-computation provides another method to estimate relative contributions of the exposure mixture components to the mixtures effect on cognitive decline (Keil et al., 2020). Quantile-based G-computation uses G-computation to estimate the total mixture effect on the outcome as a one quantile increase in all mixture components at the same time. Additionally, the weight of each exposure in the mixture effect estimate is calculated, providing the proportion of the partial effect due to a specific exposure (Keil et al., 2020). All analyses were done using the *qgcomp* package in R (ver. 3.6.1). The adjusted model was fitted for four quantiles of exposure and 500 bootstrap samples.

### Effect Modification by nSES

To investigate how associations between air pollution and cognitive decline were modified by specific nSES characteristics, we included interaction terms for the pollutant and those nSES characteristics with the most robust associations with cognitive decline in the individual air pollutant linear regression models. These models were adjusted for the same factors as the models above.

### Joint Effects

We applied the SOM algorithm in order to identify clusters of census tracts with similar air pollution and nSES characteristics (Pearce et al., 2014, 2016). The number of clusters identified by the SOM algorithm was determined by identifying group structure using within cluster sum of squares and between cluster sum of squares statistics, as well as visual inspection of the cluster star plot. These methods identify clusters with exposure levels homogenous within the cluster and heterogenous between clusters. Census tract clusters were next matched to EHAS participants using the census tract of the participant’s address. Once participants were assigned a SOM exposure cluster, adjusted linear regression models estimated the effect of exposure cluster on CFI score. The reference cluster for the linear regression model was the cluster with highest nSES and lowest air pollution concentrations. The result is a model describing the joint effect of observed mixture combinations of air pollution and nSES characteristics on CFI. All SOM analyses were performed using R version 3.6.1 (R Core Team, Vienna, Austria) using code within the *ECM* package available at: https://github.com/johnlpearce/ECM.

## Results

### Population Characteristics

The final study sample contained 11,897 individuals, 50 years and older, living in the Metro-Atlanta area. These participants had a median age of 65 years, and were majority white race (81.9%). Participants of EHAS were well educated, with 73.3% having a bachelor’s degree or higher. The median CFI score was 1.5 (IQR: 2.5) (Table 1), and the CFI distribution was right skewed.

**Table 1.**
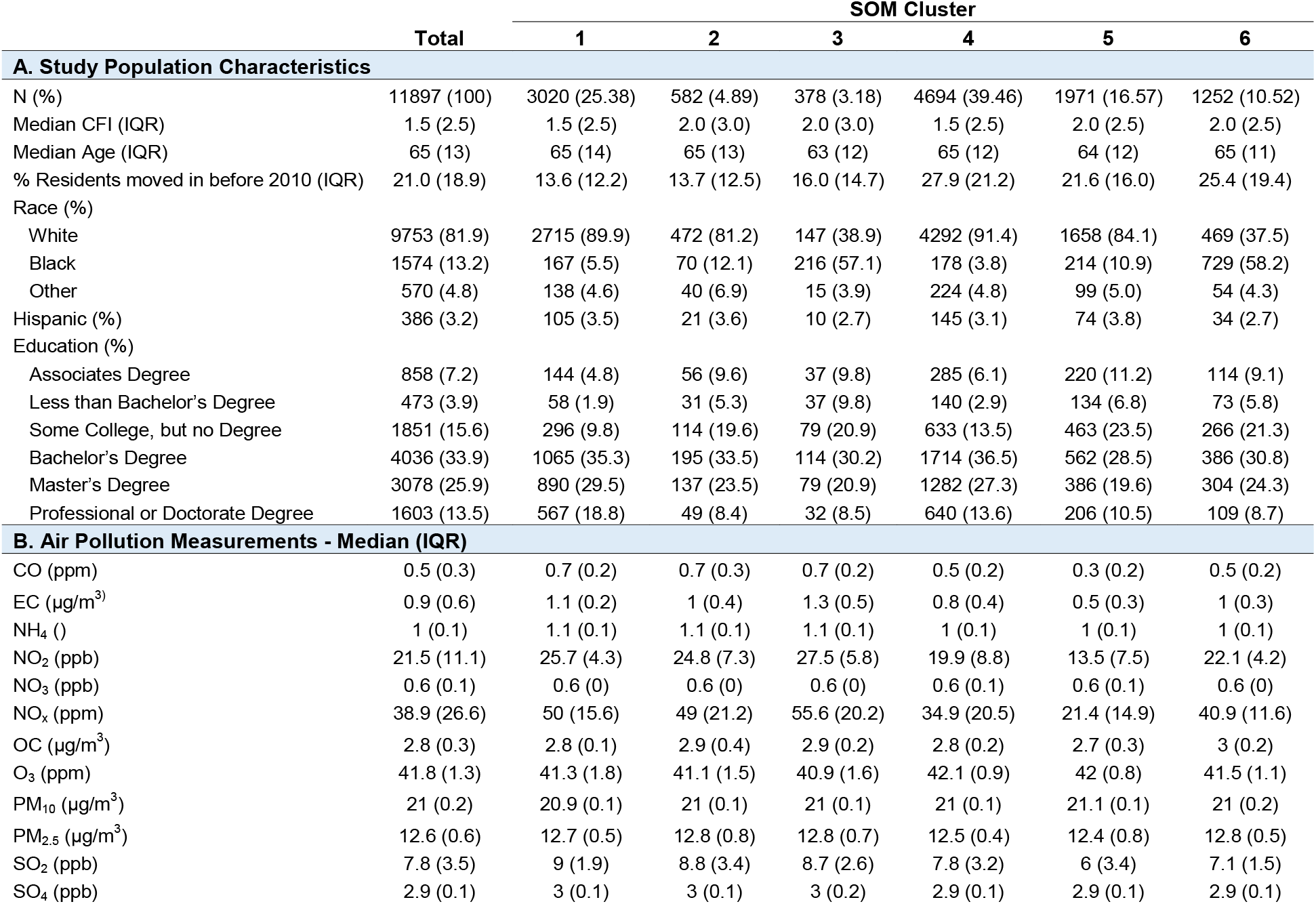
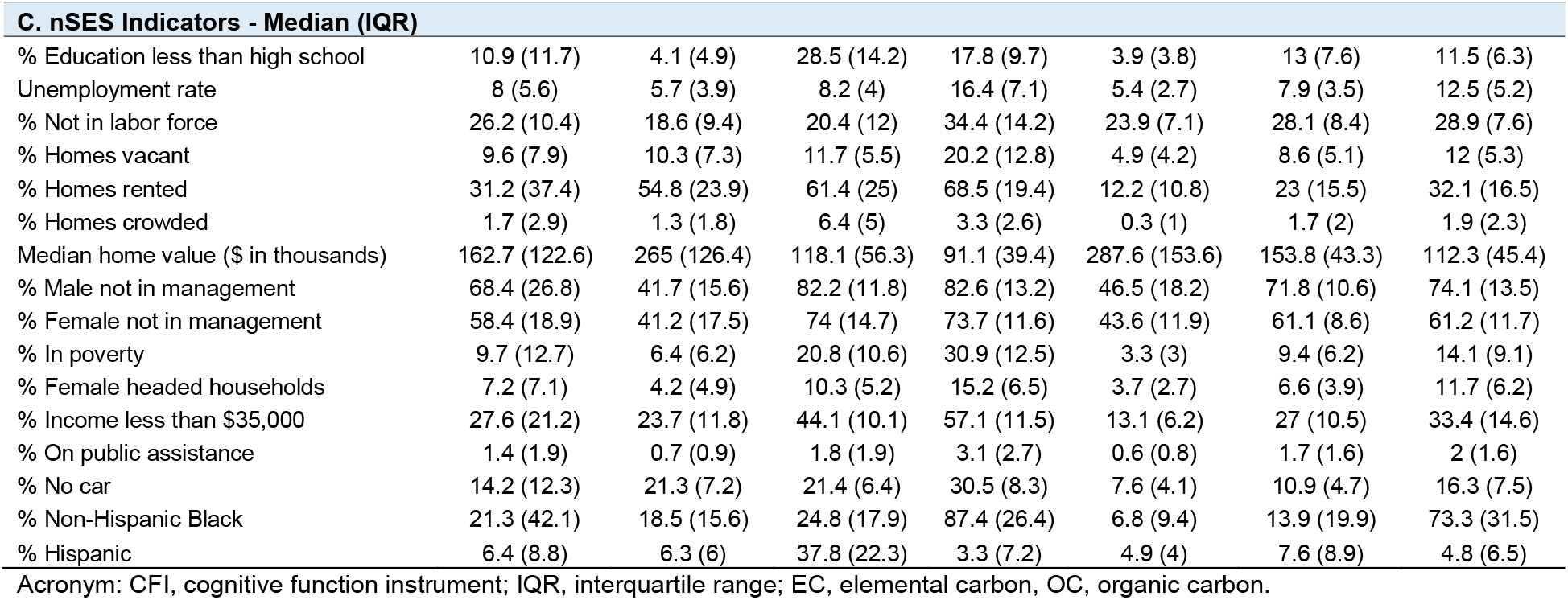
A. Emory Healthy Aging Study (EHAS) study population characteristics by Self-Organized Map (SOM) cluster. B. Air pollution measurements by SOM cluster. C. Neighborhood Socioeconomic Status (NSES) indicators by SOM cluster.

Air pollutants were positively correlated with one another, except for O_3_ and PM_10_, which had negative correlation with other air pollutants (Figure S2). Similarly, nSES characteristics were positively correlated with one another, with the exception of median home value, for which higher values correspond to higher nSES leading to a negative correlation with most other nSES characteristics. In order to make the results of the association analyses easier to interpret, we multiplied the median home value by (−1) for all subsequent analyses. This way higher values of all nSES characteristics correspond to lower nSES. Air pollutants and nSES characteristics were not highly correlated with each other, though percent with no car, and percent rented homes were positively correlated with most air pollutants, except for O_3_ and PM_10_. Average median home value had a small positive correlation with most air pollutants, except for OC, PM_10_ and PM_2.5_.

### Individual Exposure Effects

In contrast to our hypothesis, individual adjusted linear regression models showed mainly protective effects for air pollutants (Figure 1A, Supplementary Table S1). An IQR increase in any of CO, EC, NH_4_, NO_2_, NO_3_, NO_x_, OC, and PM_2.5_ each showed either null effects or significant protective effects on cognitive decline, even when adjusted for PCs of nSES characteristics and individual level confounders. The pollutant with the largest protective effect was CO, in the adjusted model an IQR increase in CO resulted in a 0.038-point decrease in ln(CFI) score (95% CI: -0.06, -0.01) indicating less cognitive decline. NO_2_ and EC had similar effects, beta estimates of -0.034 (95% CI: -0.06, -0.01) and -0.031 (95% CI: -0.05, -0.01), respectively (figure 1A, supplementary table 1).

**Figure 1.**
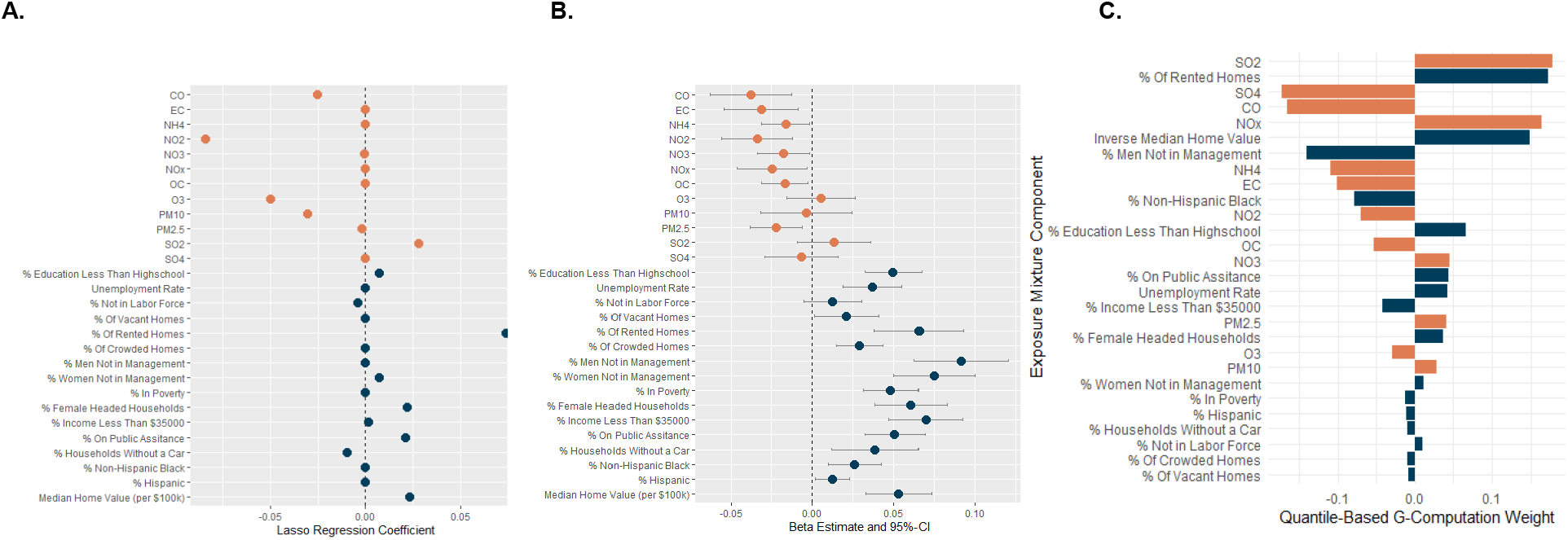
The effect of an IQR increase in air pollution and neighborhood socio-economic status (nSES) exposures on ln(CFI) score. Air pollution exposure measurements were taken from the CMAQ chemical transport model in 2008-2010, and neighborhood socio-economic status exposures are census tract averages from 2013-2018. Median home value was multiplied by -1 so that a high value was the same nSES direction as other nSES characteristics. **A**. Lasso regression coefficients of air pollutants and nSES characteristics adjusted for individual age, race/ethnicity, education, and residential stability of the census tract. **B**. Linear regression models were adjusted for individual age, race/ethnicity, education, and residential stability of the census tract, air pollution exposure models were additionally adjusted for principal components of nSES characteristics and nSES clusters. **C**. Quantile-Based g-computation weights, census tract matched air pollutant measurements. Bar lengths are comparable in magnitude among each side only. Orange bars represent air pollutants while blue bars represent nSES characteristics. **e 1**. The effect of an IQR increase in air pollution and neighborhood socio-economic status (nSES) exposures on ln(CFI) score. Air pollution ure measurements were taken from the CMAQ chemical transport model in 2008-2010, and neighborhood socio-economic status exposures ensus tract averages from 2013-2018. Median home value was multiplied by -1 so that a high value was the same nSES direction as other characteristics.

In line with our hypothesis, an IQR increase in almost all nSES characteristics (indicate lower nSES) showed significant harmful effects on cognitive decline when adjusted for individual level confounders. The magnitude of an IQR increase in any of the nSES characteristic’s effects on ln(CFI) were also relatively small, all less than 0.01. An IQR increase in percent of rented homes in the census tract resulted in a 0.066-point (95% CI: 0.04, 0.09) increase in ln(CFI) score, indicating increased cognitive decline in adjusted linear regression models. Similarly, negative median home value (median home value x (−1)) showed harmful effects per IQR increase (beta: 0.053; 95% CI: 0.03, 0.07) (figure 1A, supplementary table 1).

The adjusted LASSO regression model showed similar trends as the linear regression models (Figure 1B, Supplementary Table S1). Among the air pollutants, the strongest observed associations were found for CO (beta: -0.0252), NO2 (beta: -0.0841), O3 (beta: -0.0501), PM_10_ (beta: -0.0306), and SO_2_ (beta: 0.0282). Other effect estimates were shrunk close or exactly to zero. An IQR increase in CO, NO_2_, O_3_, PM_10_, and PM_2.5_ showed a protective effect against cognitive decline, whereas an IQR increase in SO_2_ showed harmful effects. In contrast to the linear regression models (Figure 1A), for nSES characteristics, an IQR increase in percent not in labor force, and percent of households without a car showed protective effects, though their effect estimates were shrunk close to 0. Percent of rented homes (beta: 0.074), percent of female headed households (beta: 0.0222), percent on public assistance (beta: 0.0213), and negative median home value (beta: 0.0321) showed harmful effects on cognitive decline, which is in line with the results from the linear regression analysis. Remaining estimates were shrunk close or exactly to 0 (Figure 1B, Supplementary Table S1).

Using address-based matching for CMAQ air pollutant exposures did not meaningfully change the results of the linear or LASSO regressions (Figure S3). For example, NO_2_ had the strongest observed association with cognitive decline in both the census tract and address matched LASSO models (Beta:-0.0841 vs. -0.0882, respectively; Table S1.) In the linear regression models, census tract matched exposures were similar to address matched exposures. For example, the effect estimate of an IQR increase in CO was -0.038 in the census tract matched model, compared to -0.04 in the address matched model (Table S1). Given these results, we proceeded with the census tract matched models to make exposure assessment comparable across models.

Using the adjusted BKMR model, individual effects of the mixture components on ln(CFI) score can be determined, while controlling for co-exposures and confounding. Figure 2A shows the individual effects of each mixture component on ln(CFI) score with all other mixture components are fixed to the 50th percentile, adjusted for confounders. Most mixture components had null estimated effects on ln(CFI) score. CO was negatively associated with the ln(CFI) score and SO_2_, percent of rented homes, and negative median home value in the census tract were positively associated with ln(CFI), which is in line with the linear regression and LASSO regression results reported above.

**Figure 2.**
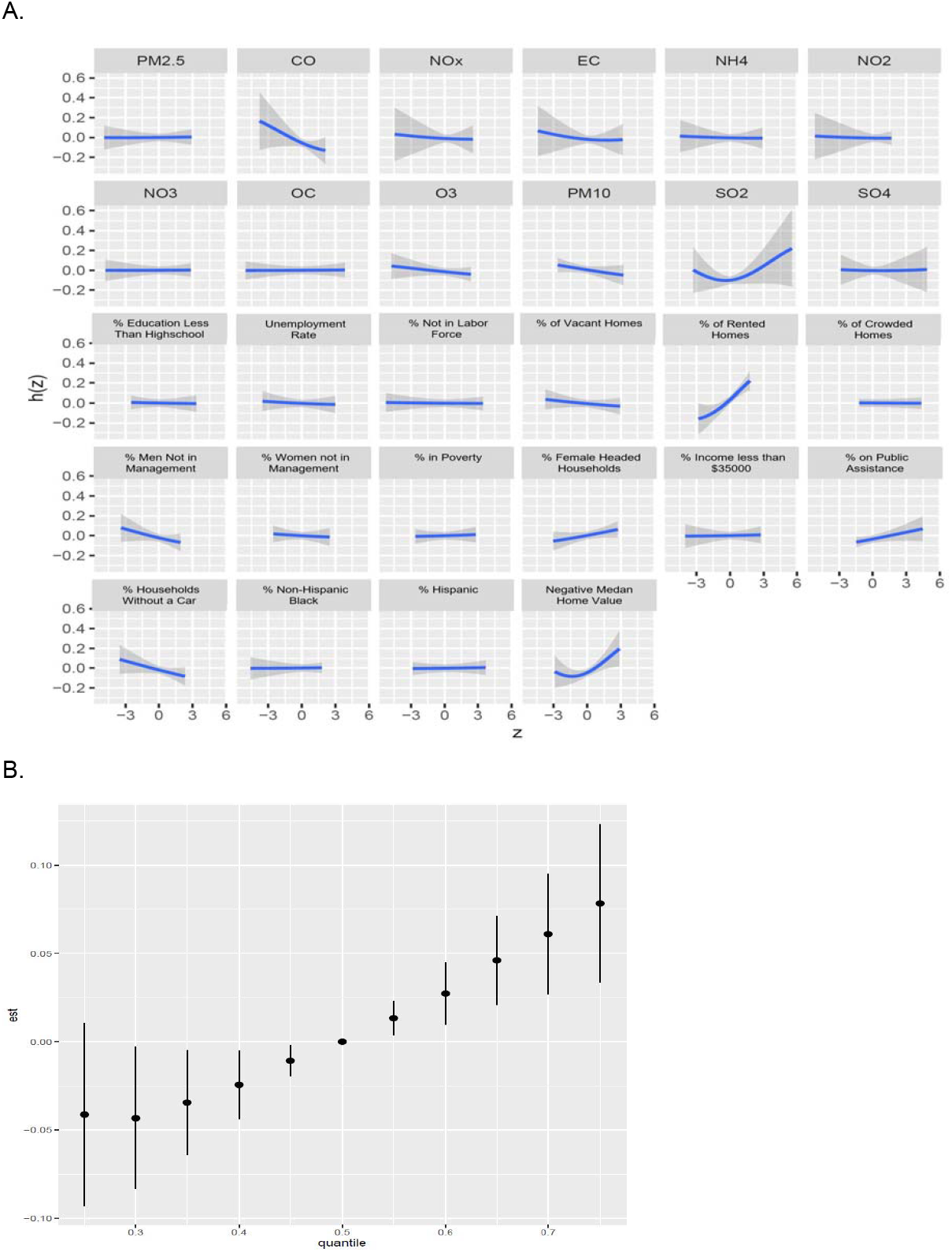
A. BKMR exposure mixture individual effects plot. The effect of each exposure holding all other exposures at their median and controlling for confounders. The y-axis for each plot represents the exposure response function (h), while the x-axis represents the centered exposure level. B. BKMR exposure mixture overall effects plot. The effect of all exposures in the mixture at each quantile compared to the effect of all exposures in the mixture at the median.

Quantile-based G-computation results suggest a lack of directional homogeneity with respect to air pollutant and nSES characteristic mixture component effects, agreeing with the linear regression analysis and the lasso regression results. In Figure 1C positive weights indicate a positive, or harmful, effect of the mixture component on ln(CFI) score. Conversely, a negative weight would indicate a protective effect of the mixture component on ln(CFI) score. In the adjusted model, SO_2_ (weight: 0.1658), and percent of rented homes (weight: 0.1726) had the most harmful estimated effects, which is in line with the results of the single pollutant models and BKMR. Conversely, SO_4_ and CO had the most protective estimated effects in adjusted models. However, the total mixture effect was not statistically significant (psi: 0.074; p-value: 0.062).

Across methods, CO, NO_2_, and O_3_ showed small protective estimated effects against increased CFI scores, while SO_2_ was the only air pollutant that had a consistently harmful estimated effect on CFI score. For the nSES characteristics, all nSES indicators had harmful estimated effects across models, particularly, percent of rented homes and negative average median home value of the census tract.

To explore potential explanations for the protective effects of air pollutants on ln(CFI) which we found in the individual exposure models, we investigated interactions and joint effects between exposures.

### Effect Modifications and Joint Exposure Effects

We analyzed whether associations between air pollution and cognitive decline were modified by nSES using linear regression models with an interaction term. Percent of rented homes and negative median home value were selected as nSES characteristics for the effect modification analyses because they showed the strongest associations with CFI in the BKMR model (Figure 2A, Table S3). We found significant estimated effect modification by percent of rented homes for the association between EC and ln(CFI) score (interaction estimate: 0.058; 95% CI: 0.015, 0.101) (Figure S4A); suggesting that the effects of EC and high percent rented homes are synergistic and ln(CFI) score is higher (increase in cognitive decline) for those with both high EC and high percent rented homes in the census tract. Additionally, the estimated effect modification by negative median home value was significant for the association between PM_10_ and ln(CFI) score (interaction estimate: 0.063; 95% CI: 0.015, 0.110) was significant, showing that high PM_10_ and low home values act together in increasing the ln(CFI) score (Figure S4B). Associations with other air pollutants were not significantly modified by percent rented homes or median home value (Table S3).

Next, we investigated the joint effects of air pollution and nSES using BKMR, quantile-based G-computation and SOM. BKMR analysis showed significant overall mixture effects (Figure 2B). The ln(CFI) score showed a significant increase when all mixture components were above their median compared to when all mixture components were at the median level. This trend suggests a linearly increasing effect estimate at all quantiles of the exposure mixture above the median. Additionally, there is also a significant decrease in the overall mixture effect on ln(CFI) when mixture components are below the median, compared to when all mixture components are at the median level. However, the 95% credible interval for the 25^th^ percentile of exposure compared to the median is not significant. Similar to earlier results, quantile-based G-computation also found a positive association between increasing values of the mixture and ln(CFI), though the total mixture effect was not statistically significant (psi: 0.074; p-value: 0.062).

Next, we used the SOM approach to investigate observed patterns of exposure mixtures in association with ln(CFI). The SOM identified 6 clusters (Figure 3A, Table 1), based on the air pollution and nSES characteristics of the census tracts. Participants were not evenly distributed across clusters, a majority of participants experiencing pollution and nSES values within in cluster 4 (n=4,694; 39.46%), while comparatively few experience the combinations of pollutants and nSES variables found in clusters 2 (n=582; 4.89%) and 3 (n=378; 3.18%). All clusters had similar median ages. Cluster 4 has the lowest concentrations of most air pollutants and highest nSES, though it has highest O_3_ and PM_10_. Conversely, cluster 3 has the highest concentrations for most air pollutants and lowest nSES. In the map of Metro-Atlanta (Figure 3B), census tracts are color coded based on their SOM cluster assignment. Cluster 4, the low pollution and high nSES cluster, seen on the map in black, primarily appears in the northern half of Metro-Atlanta outside of the City of Atlanta. Clusters 1, 2, and 3 appear primarily in the City of Atlanta around highways (black lines on map). These three clusters also have the highest proportions of rented homes. In line with Atlanta’s historical segregation policies, clusters 3 and 6, located in southern Atlanta, contain census tracts with the highest proportions of non-Hispanic Black residents. A majority of Black EHAS participants live in cluster 6.

**Figure 3.**
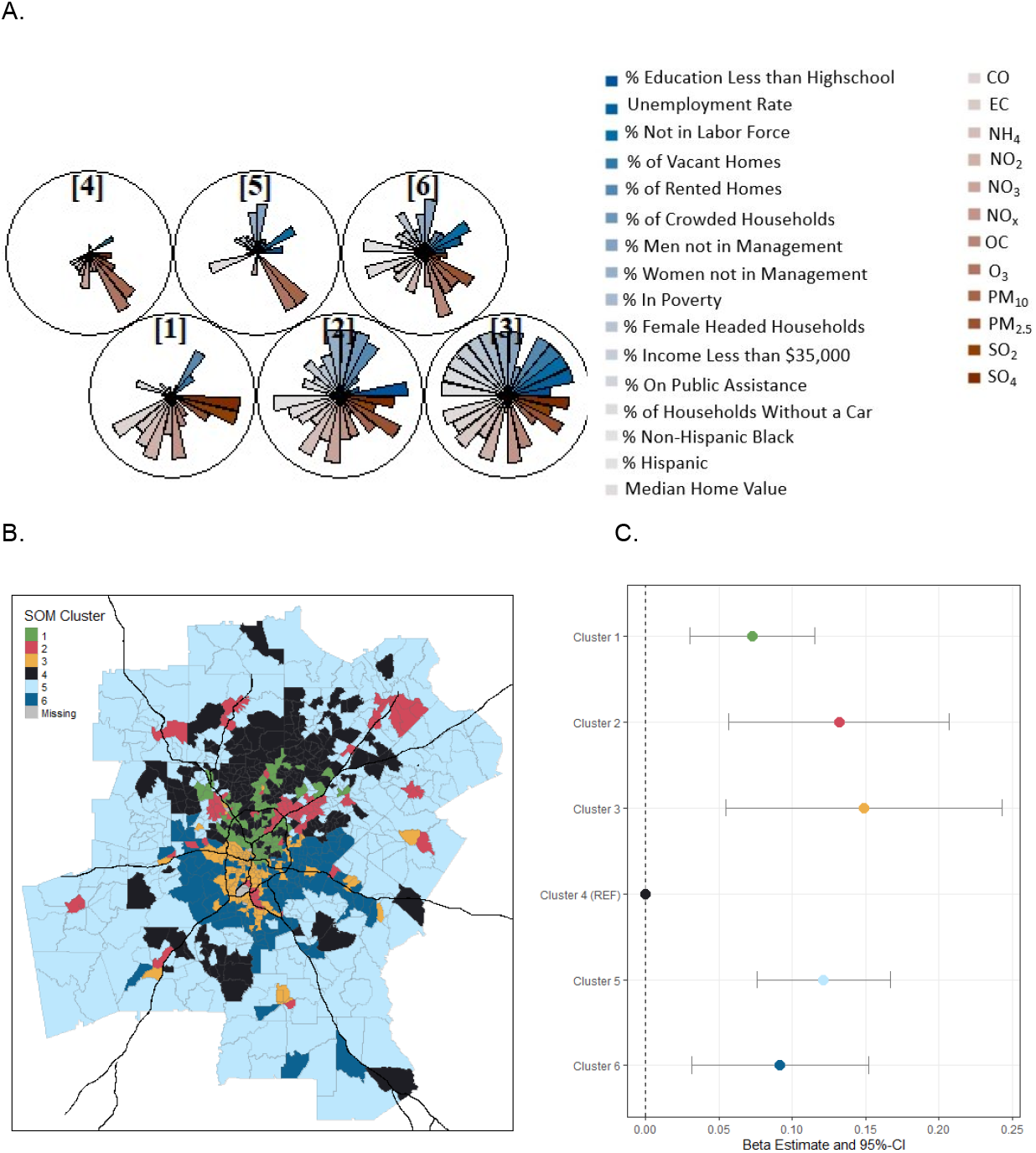
Associations between air pollution and neighborhood socio-economic status (nSES) indicators for cognitive functioning. Median home value was multiplied by -1 so that a high value was the same nSES direction as other nSES characteristics **A**. SOM cluster star plot, slices represent median values of a mixture component, each circle is a SOM cluster. Blue slices correspond with nSES indicators, while red slices correspond with air pollutants **B**. Map of census tracts in Metro-Atlanta by cluster. Black lines represent major highways. **C**. Results of linear regression model estimating SOM cluster effect on ln(CFI) score. Cluster 4, the highest nSES cluster, was used as the reference group. Model adjusted for individual age, race/ethnicity, education, and residential stability of the census tract.

In the adjusted linear regression model using indicators of SOM cluster membership as exposure, cluster 4 was used as the reference group because it had the lowest air pollution and highest nSES. Compared to cluster 4, all other clusters exhibit increased ln(CFI) scores, indicating increased cognitive decline (Figure 3C, Supplementary Table S2). Clusters 2 and 3, which have the highest air pollution concentrations and lowest nSES, also have the highest increase in ln(CFI) compared to cluster 4, adjusting for individual age, race/ethnicity, and residential stability of the census tract. These results provide effect estimates based on observed contrasts in exposure mixtures and suggest synergistic effects of air pollution and nSES. These findings are in line with interaction results reported earlier.

The high O_3_ concentrations in cluster 4, the highest nSES cluster and cluster with the largest number of participants (39.46%), may be driving the estimated protective effect of O_3_ observed in the single pollutant models (Figure 3) because high SES potentially protects participants from the harmful effects of air pollution. NO_2_ and CO concentrations, which are highly correlated (Figure 3), are highest in clusters 1, 2, and 3, all located along highways (Figure 2B). Cluster 1 is the second largest cluster with 25.38% of participants and has the second highest nSES. The large proportion of participants in cluster 1, compared to clusters 2 (4.89%) and 3 (3.18%) may drive the estimated protective effects of NO_2_ and CO (Table 1) observed throughout the results in this analysis. Individuals within cluster 1 have higher SES that can potentially protect them from the harmful effects of air pollution, while participants with low SES in clusters 2 and 3 do not have that same protection.

## Discussion

In this study of 11,897 individuals 50 years and older from Metro Atlanta, we observed significant joint effects of air pollution and nSES on cognitive decline. Using analysis techniques accounting for multiple exposures, we disentangle the seemingly protective estimated effects of air pollution found in the individual exposure models. Air pollution and nSES exposure profiles were generated using the joint SOM model report estimated significant associations between exposure profile and cognitive decline. These associations between exposure profiles and cognitive decline yield evidence of the harmful joint effect of lower nSES and air pollution. Additionally, the BKMR model provided insight into which air pollutants and nSES characteristics most influence cognitive decline, as well as estimates of synergistic effects of the exposure mixture. These analyses reveal how imperative interaction and mixture analyses are to gain a more accurate and holistic picture of how air pollution and nSES jointly affect cognitive decline.

Previous research has shown harmful associations between air pollutants and cognitive decline (Peters et al., 2019); as well as harmful associations between nSES and cognitive decline (Besser et al., 2017; Luo et al., 2019; Steptoe & Zaninotto, 2020). While nSES and air pollution may affect cognitive decline separately, it is difficult to separate their effects because they are often highly correlated. A global review found that in North America lower SES communities experience higher levels of air pollution (Hajat et al., 2015). These correlations between air pollution and nSES are frequently observed in the environmental justice literature. However, previous epidemiologic studies have mainly focused on analyzing individual exposures without controlling for co-exposures to other environmental and social stressors. In diverse cities like Atlanta, where gentrification, high levels of car traffic, and other factors often lead to high nSES neighborhoods with high air pollution, the relationship between air pollution and nSES can be more complicated. Ignoring these complicated relationships can lead to the seemingly protective effects of air pollution on cognitive decline observed in the simpler, individual air pollutant analyses. We see this complicated relationship in the overall low correlation between air pollutants and nSES characteristics, with only the nSES characteristics percent with no car and percent rented homes being positively correlated with most air pollutants, except O_3_ and PM_10_.

To the best of our knowledge, this paper is the first to use cutting edge environmental mixture methods to study air pollution and nSES characteristics in relation to cognitive decline. Previous studies of the effect of air pollution on cognitive decline have treated nSES as a confounder or effect modifier, not a co-exposure (J. Ailshire et al., 2017; J. A. Ailshire & Crimmins, 2014; Bowe et al., 2019; Cullen et al., 2018; Li et al., 2021). Utilizing multiple mixtures method approaches such as, SOM, BKMR, and quantile-based G-computation supports a comprehensive examination of the relationship between the exposure mixtures and outcomes (Taylor et al., 2016). The SOM analysis revealed that participants in census tract clusters with high air pollution and low nSES had the largest decline in cognitive function, compared to census tracts with lower air pollution or higher nSES. SOM analyses, and other clustering methods, can lend insight into joint effects of exposure mixtures by identifying real-world contrasts to examine. Our results build upon those of another study analyzing EHAS data in Metro Atlanta, which identifies nSES as an effect modifier of the association between air pollution and cognitive decline suggesting a significantly harmful association between air pollutant levels and cognitive decline in low nSES neighborhoods (Li et al., 2021).

BKMR analysis also reveals the overall effect of increasing the mixture of high air pollution exposure and low nSES (nSES characteristics are coded to be increasing value is lower SES) significantly increases ln(CFI) score. While the quantile-based G-computation analysis did not show a significant association between the air pollution and nSES exposure mixture and ln(CFI) score, this could be due to non-linearity of the association between the exposure mixture and the outcome. The quantile-based G-computation method can handle non-linear and non-additive exposure effects, but non-linear terms need to be pre-defined by the user (Keil et al., 2020), in contrast to the data-driven investigation of non-linearity in BKMR, which does not require a priori knowledge of the relationship. BKMR can handle non-linear associations in the exposure/outcome relationship without specifically adding non-linear terms into the model (Bobb et al., 2015, 2018). The BKMR analysis illustrates that SO_2_, percent rented homes, and negative median home value have non-linear relationships with ln(CFI) score (Figure 3A). Future studies using these methods can use the results from BKMR to inform modeling using other methods.

Our results illustrate the importance of analyzing air pollution and socioeconomic exposures as a mixture. In our study, single air pollutant linear regression models estimate protective effects of air pollution exposure, counter to biological plausibility. Using methods that examine effect modifications and joint effects in linear, non-linear and spatial analyses, it was possible to disentangle the more nuanced and complicated underlying relationships between exposure to air pollution and nSES and cognitive decline in Metro Atlanta. Additionally, using multiple methods to model the effect of the exposure mixture on the outcome allowed for analysis of different aspects of the relationship between the exposure mixture and cognitive decline (Taylor et al., 2016).

A strength of this study is its large sample size from a diverse city. The large sample size of the EHAS cohort and its spatial distribution across Metro Atlanta allowed for higher precision and power than smaller studies, especially in mixture modeling analyses. The diversity of Atlanta also allowed the SOM clustering algorithm to define a variety of profiles of the exposure mixture. An additional strength of our results is the use of modeling techniques that examine individual and joint effects of air pollutants and nSES characteristics. Use of the CFI to determine cognitive decline strengthens the impact of this analysis because subjective cognitive decline is considered to be one the of first signs of progression to dementia (Amariglio et al., 2015; Jessen, 2014).

The interpretation of the results above is limited by the cross-sectional nature of our data. While air pollution measurements are from 2008-2010 which is before the EHAS enrollment window of 2015-2020, participants were asked their address to connect to the air pollution and nSES at the same time as the outcome assessment. There is possible exposure misclassification based on participants moving prior to their enrollment in EHAS, though this is likely to be non-differential by CFI score. An additional limitation in the exposure assessment is the grid size of the CMAQ chemical transport model. The CMAQ chemical transport model has 4 km resolution grids, which may not capture all of the variation in air pollution levels when looking at Metro-Atlanta.

However, another study using air pollution exposure models with a 250m resolution did report similar harmful effects of air pollutants on cognitive decline (Li et al., 2021). Additionally, EHAS is not a population-based sample, and the majority of participants are White and live in high nSES areas and are of high SES individually which is not the case in the Atlanta general population. There may be limitations to the generalizability of these results.

Our results reveal how imperative it is to include nSES into analyses of air pollution and cognitive decline, not just as a confounder of environmental pollution levels but as a co-exposure with environmental pollution. Future studies examining such co-exposures should include longitudinal measurements of air pollution and cognition to move further toward causal inference for these joint associations.

## Data Availability

Data requests are handled through the Emory University Goizueta Alzheimer's Disease Research Center (https://alzheimers.emory.edu/research/for-researchers/data-request-form.html).

## Acknowledgements

Research reported in this publication was supported by the National Institute of Environmental Health under Award Number 5T32ES12870. The content is solely the responsibility of the authors and does not necessarily represent the official views of the National Institutes of Health.

This work was based on information from the Emory Healthy Aging study, supported by HERCULES Pilot Project via National Institute of Environmental Health Sciences (NIEHS) P30ES019776 (PI: Anke Huels) and National Institute on Aging (NIA) R01AG070937 (PI: James J. Lah).

