## Supplemental Materials for "The complex relationship of air pollution and neighborhood socioeconomic deprivation and their association with cognitive decline"

**Supplementary Material**

**Joint effects of air pollution mixtures and neighborhood socioeconomic deprivation on cognitive decline in Metro Atlanta, USA**

Grace Christensen^1^, Zhenjiang Li^2^, John Pearce^3^, Michele Marcus^1,2^, James J. Lah^4^, Lance A. Waller^2,5^, Stefanie Ebelt^1,2^, Anke Huels^1,2^

^1^ Department of Epidemiology, Rollins School of Public Health, Emory University, Atlanta, GA, USA,

^2^ Gangarosa Department of Environmental Health, Rollins School of Public Health, Emory University, Atlanta, GA, USA

^3^ Department of Public Health Sciences, College of Medicine, Medical University of South Carolina, Charleston, SC, USA

^4^ Department of Neurology, School of Medicine, Emory University, Atlanta, GA, USA

^5^ Department of Biostatistics and Bioinformatics, Rollins School of Public Health, Emory University, Atlanta, GA, USA

**Corresponding author:**

Anke Huels, PhD

Rollins School of Public Health, Emory University

1518 Clifton Road, Atlanta, GA 30322

**Table S1.** Regression coefficients for Lasso and linear regressions using both census tract (CT) and participant address matchs air pollutant exposures. Additionally, quantile-based g-computation weights for all exposures based on CT matched air pollution concentrations.

|  | Lasso Regression Coefficient (CT match) ** | Lasso Regression Coefficient (address match)** | Linear Regression Beta (95% CI) (CT match) ** | Linear Regression Beta (95% CI) (address match) ** | Quantile-Based G-Computation Weight (CT match)** |
| --- | --- | --- | --- | --- | --- |
| CO (ppm) | -0.0252 | -0.0131 | -0.038 (-0.06,-0.01) | -0.04 (-0.07,-0.01) | -0.1658 |
| EC (μg/m^3)^ | . | . | -0.031 (-0.05,-0.01) | -0.036 (-0.06,-0.01) | -0.1014 |
| NH_4_ | . | -0.0020 | -0.016 (-0.03,0) | -0.018 (-0.03,0) | -0.1088 |
| NO_2_ (ppb) | -0.0841 | -0.0882 | -0.034 (-0.06,-0.01) | -0.036 (-0.06,-0.01) | -0.0694 |
| NO_3_ (ppb) | -0.0006 | . | -0.018 (-0.03,0) | -0.019 (-0.03,0) | 0.0457 |
| NO_x_ (ppm) | . | . | -0.025 (-0.05,0) | -0.029 (-0.05,-0.01) | 0.1644 |
| OC (μg/m^3^) | . | -0.0030 | -0.017 (-0.03,0) | -0.016 (-0.03,0) | -0.0538 |
| O_3_ (ppm) | -0.0501 | -0.0508 | 0.006 (-0.02,0.03) | 0.01 (-0.01,0.03) | -0.0292 |
| PM_10_ (μg/m^3^) | -0.0306 | -0.0342 | -0.004 (-0.03,0.02) | -0.006 (-0.03,0.02) | 0.0279 |
| PM_2.5_ (μg/m^3^) | -0.0018 | . | -0.022 (-0.04,-0.01) | -0.023 (-0.04,-0.01) | 0.0412 |
| SO_2_ | 0.0282 | 0.0444 | 0.014 (-0.01,0.04) | 0.011 (-0.01,0.03) | 0.1781 |
| SO_4_ | . | -0.0249 | -0.007 (-0.03,0.02) | -0.012 (-0.04,0.01) | -0.1717 |
| % Education less than high school | 0.0074 | 0.0114 | 0.05 (0.03,0.07) | 0.05 (0.03,0.07) | 0.0659 |
| Unemployment rate | . | . | 0.037 (0.02,0.06) | 0.037 (0.02,0.06) | 0.0424 |
| % Not in labor force | -0.0038 | -0.0078 | 0.013 (-0.01,0.03) | 0.013 (-0.01,0.03) | 0.0098 |
| % Homes vacant | . | . | 0.021 (0,0.04) | 0.021 (0,0.04) | -0.0085 |
| % Homes rented | 0.0740 | 0.0904 | 0.066 (0.04,0.09) | 0.066 (0.04,0.09) | 0.1726 |
| % Homes crowded | . | -0.0029 | 0.029 (0.01,0.04) | 0.029 (0.01,0.04) | -0.0096 |
| % Male not in management | . | . | 0.092 (0.06,0.12) | 0.092 (0.06,0.12) | -0.1399 |
| % Female not in management | 0.0071 | 0.0058 | 0.075 (0.05,0.1) | 0.075 (0.05,0.1) | 0.0121 |
| % In poverty | . | . | 0.048 (0.03,0.07) | 0.048 (0.03,0.07) | -0.0117 |
| % Female headed households | 0.0222 | 0.0202 | 0.061 (0.04,0.08) | 0.061 (0.04,0.08) | 0.0367 |
| % Income less than $35,000 | 0.0014 | 0.0079 | 0.07 (0.05,0.09) | 0.07 (0.05,0.09) | -0.0422 |
| % On public assistance | 0.0213 | 0.0211 | 0.051 (0.03,0.07) | 0.051 (0.03,0.07) | 0.0431 |
| % No car | -0.0098 | -0.0317 | 0.039 (0.01,0.07) | 0.039 (0.01,0.07) | -0.0100 |
| % Non-Hispanic Black | . | . | 0.026 (0.01,0.04) | 0.026 (0.01,0.04) | -0.0788 |
| % Hispanic | . | -0.0015 | 0.013 (0,0.02) | 0.013 (0,0.02) | -0.0110 |
| Median home value (per $100k)* | 0.0231 | 0.0262 | 0.053 (0.03,0.07) | 0.065 (0.04,0.09) | 0.1486 |
| * median home value was multiplied by -1 to go in the direction of the other nSES variables | | | | | |
| ** adjusted for individual age, race/ethincity, education, and residential stability. In linear regression models with air pollution exposure models were additionally adjusted for principal components of nSES characteristics and nSES clusters | | | | | |
| CT = census tract | | | | | |

**Table S2.** Joint effects of air pollution and nSES on subjective cognitive decline (CFI score). Linear regression models of air pollution and nSES on CFI score, using SOM cluster as the exposure. Adjusted model adjusted for individual age, race/ethnicity, education, and residential stability. Exposure clusters are detailed in Figure 2.

|  | Beta Estimate (95% CI) | |
| --- | --- | --- |
|  | Crude | Adjusted |
| Cluster 1 | 0.036 (-0.003, 0.074) | 0.073 (0.030, 0.115) |
| Cluster 2 | 0.146 (0.072, 0.219) | 0.132 (0.057, 0.207) |
| Cluster 3 | 0.209 (0.120, 0.298) | 0.149 (0.055, 0.243) |
| Cluster 4 | REF | REF |
| Cluster 5 | 0.153 (0.108, 0.197) | 0.121 (0.076, 0.167) |
| Cluster 6 | 0.160 (0.107, 0.213) | 0.092 (0.032, 0.152) |

**Table S3. Effect modification of the relationship between air pollutants and CFI score by select nSES characteristics.** Individual air pollutant linear regression models with interaction term for the air pollutant and nSES characteristic. Beta estimate and 95% CI’s are shown for interaction terms. Models adjusted for individual age, race/ethnicity, education, and residential stability

|  | **Percent of homes rented** | **Negative median home value** |
| --- | --- | --- |
| **Pollutant** |  |  |
| CO | 0.029 (-0.019, 0.076) | -0.003 (-0.042, 0.037) |
| EC | **0.058 (0.015, 0.101)** | 0.007 (-0.03, 0.044) |
| NH_4_ | 0.009 (-0.024, 0.042) | 0.011 (-0.017, 0.039) |
| NO_2_ | 0.033 (-0.01, 0.077) | -0.001 (-0.037, 0.035) |
| NO_3_ | 0.006 (-0.034, 0.046) | 0.009 (-0.026, 0.044) |
| NO_x_ | 0.033 (-0.008, 0.074) | 0 (-0.034, 0.034) |
| OC | 0.029 (-0.001, 0.059) | 0.004 (-0.019, 0.027) |
| O_3_ | -0.017 (-0.058, 0.025) | -0.012 (-0.043, 0.019) |
| PM_10_ | -0.022 (-0.076, 0.032) | **0.063 (0.015, 0.11)** |
| PM_2.5_ | 0.029 (-0.005, 0.063) | 0.008 (-0.016, 0.032) |
| SO_2_ | 0.011 (-0.035, 0.057) | 0.023 (-0.019, 0.066) |
| SO_4_ | 0.027 (-0.017, 0.071) | 0.028 (-0.014, 0.069) |
| **bold:** significant interaction at alpha=5% significance level | | |

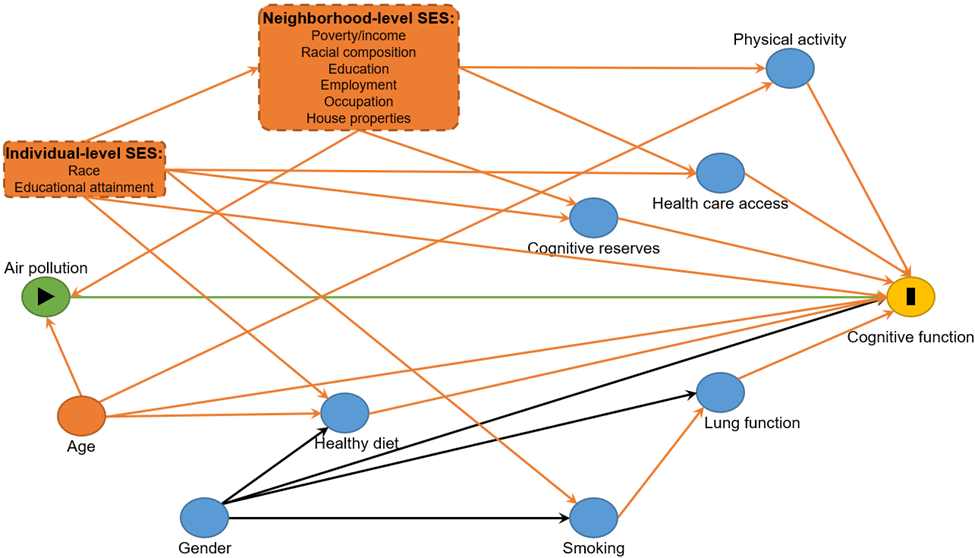

**Figure S1.** Directed Acyclic Graph (DAG) of the hypothesized causal structure linking air pollution, neighborhood level SES, and cognitive function. The biasing paths and the minimal sufficient set of confounders for identifying the total effect of ambient air pollution on cognitive function are colored in orange.

A.	B.

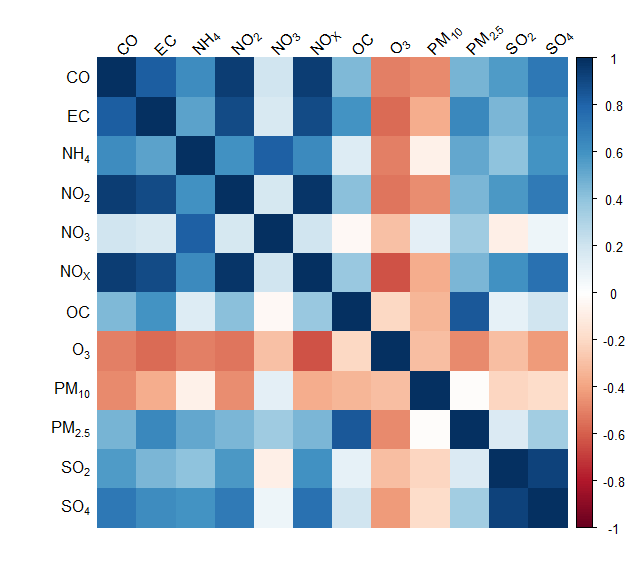

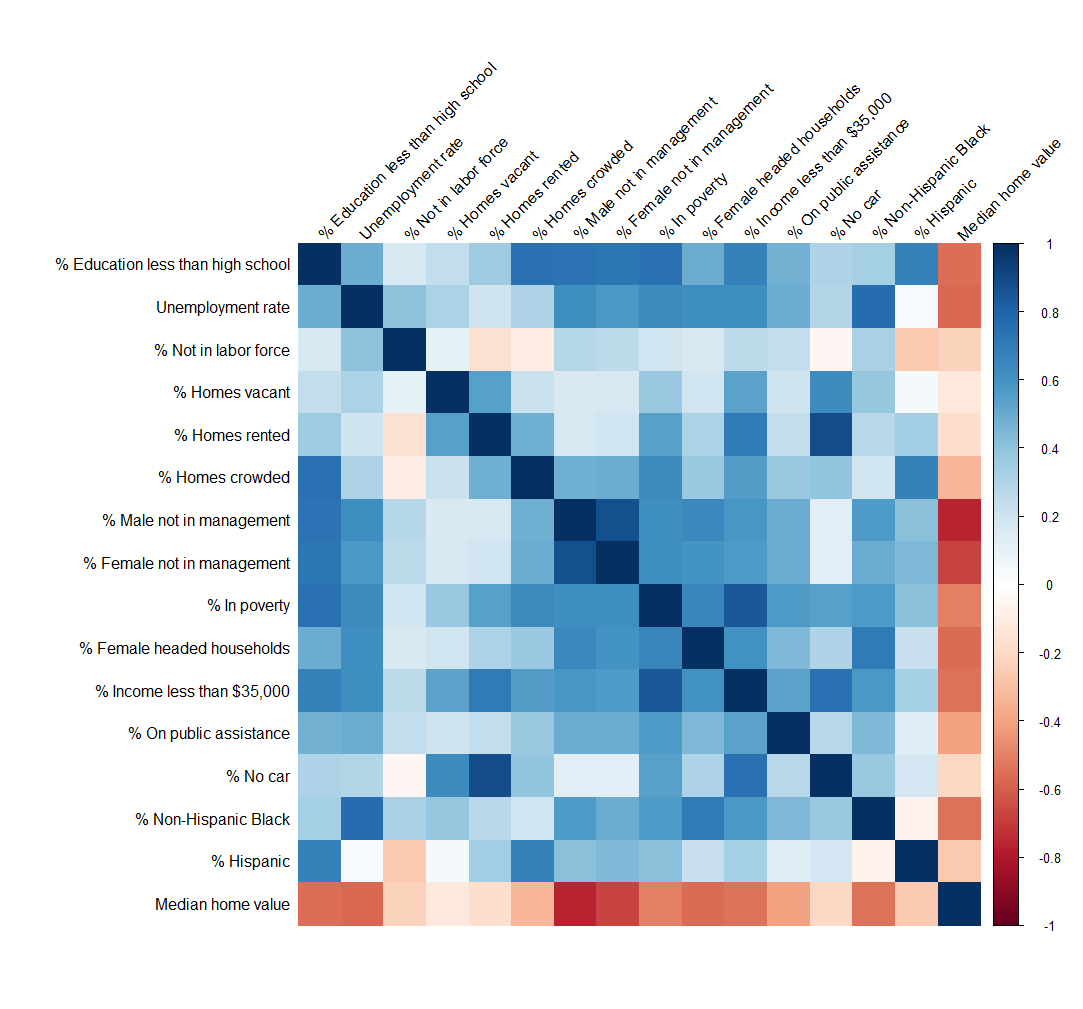

C.

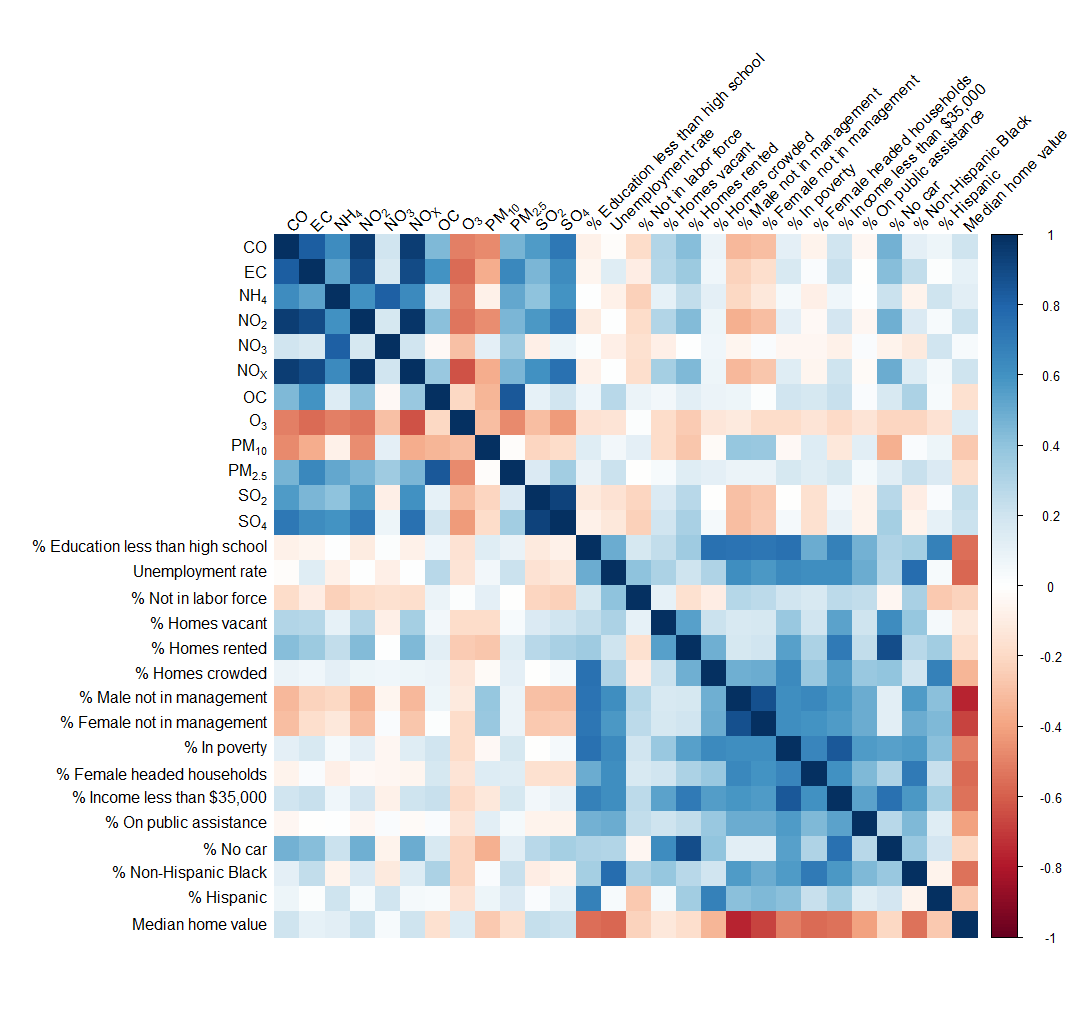

**Figure S2.** Correlation matrix of air pollutants and neighborhood level socioeconomic status (nSES) characteristics. Median home value has not been manipulated. Air pollutants are census tract matched.

**A.	B.**

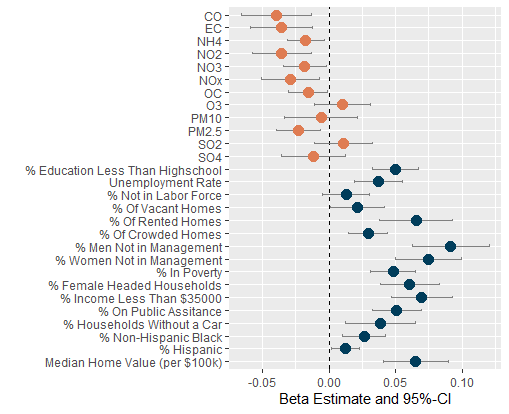

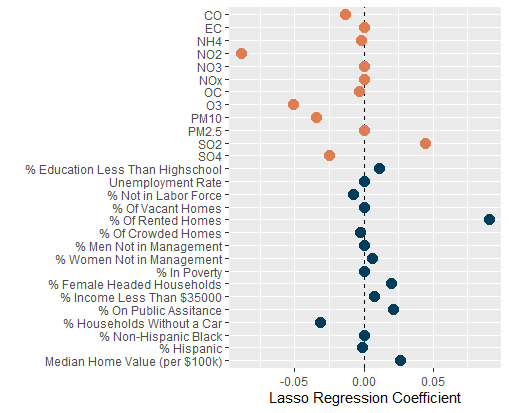

**Figure S3.** The effect of an IQR increase in air pollution and neighborhood socio-economic status (nSES) exposures on lnCFI score. Air pollution exposure measurements were taken from the CMAQ chemical transport model in 2008-2010, and neighborhood socio-economic status exposures are census tract averages from 2013-2018. CMAQ air pollution exposure concentrations were matched to participants based on the address given at enrollment.Median home value was multiplied by -1 so that a high value was the same nSES direction as other nSES characteristics. **A.** Linear regression models were adjusted for individual age, race/ethnicity, education, and residential stability of the census tract, air pollution exposure models were additionally adjusted for principal components of nSES characteristics and nSES clusters. **B.** Lasso regression coefficients of air pollutants and nSES characteristics adjusted for individual age, race/ethnicity, education, and residential stability of the census tract.

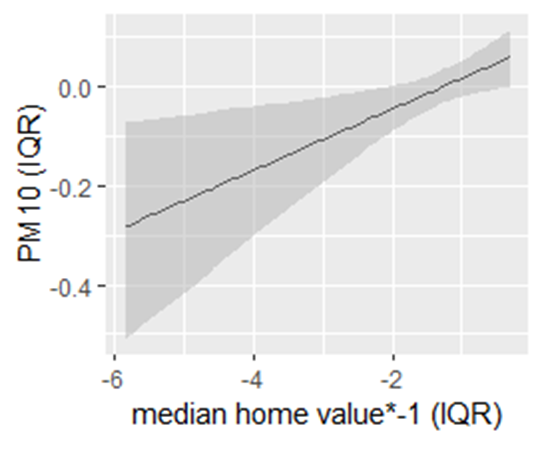

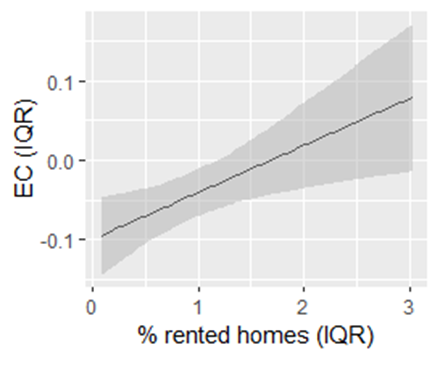
A.	B.

**Figure S4.** Statistically significant effect modification of individual air pollutants and nSES characteristics on cognitive decline. Linear regression models adjusted for individual age, race/ethnicity, education, and residential stability **A.** The effect estimate of EC on cognitive decline increases within increased percentage of rented home. **B.** The effect estimate of PM10 on cognitive decline increases with decreasing median home value.
